# Sleep as a Modifiable Risk Factor for Childhood Autism: Stratified Analysis of U.S. National Survey of Children’s Health Data

**DOI:** 10.1101/2025.08.12.25333516

**Authors:** Md Roungu Ahmmad, Harry Pantazopoulos, Fazlay Faruque, Xiaoli Zhang, Reecha Puri

## Abstract

**Purpose:** This study aimed to examine the association between age-specific sleep sufficiency and autism spectrum disorders (ASD) among U.S. children aged 6–17 years.

**Methods:** Data were gathered from the 2022-2023 National Survey of Children’s Health (NSCH), including 63,866 children. Sleep sufficiency was defined based on age-specific guidelines from the American Academy of Sleep Medicine. Descriptive statistics, incidence risk ratios (IRRs), and adjusted logistic regression models were used to assess associations between ASD and key predictors. Stratified models by sex and BMI were conducted to explore effect modification. Additionally, a machine learning model was developed to predict the adjusted probability of ASD risk.

**Results:** Children with insufficient sleep had a significantly higher incidence of ASD (5.16%) compared to those with sufficient sleep (4.05%) (p < 0.001). In adjusted models, sufficient sleep was associated with lower odds of ASD (OR = 0.78; 95% CI: 0.72–0.85; p < 0.001). Stratified analyses showed a protective effect in both males (OR = 0.78; 95% CI: 0.71–0.86) and females (OR = 0.80; 95% CI: 0.68–0.93), more pronounced in males. Machine learning analysis revealed that females with sufficient sleep and age below 14 years exhibited the lowest probability of ASD, whereas males aged 8 to 14 years with insufficient sleep demonstrated the highest likelihood of ASD risk.

**Conclusion:** These results suggest that sufficient age-specific sleep is significantly associated with reduced odds of ASD, particularly in male children. Findings highlight the importance of sleep as a potentially modifiable factor in ASD risk and support targeted public health interventions.

## Background

Autism Spectrum Disorder (ASD) is a complex neurodevelopmental condition characterized by impairments, and deficits in social communication, as well as the presence of restricted, and repetitive behaviors or interests (Hirota & King, 2023). The prevalence of ASD is 3 to 4 times greater in males than in females (Hirota & King, 2023; Maenner, 2021, 2023). In recent decades, the diagnosis of ASD has substantially increased in the United States, one in 44 children in 2018 and one in 36 in 2020 (Maenner, 2021, 2023). Early diagnosis of ASD can have a significant positive impact (Dawson et al., 2010; Kasari et al., 2010), providing potential opportunity for early intervention strategies. Therefore, it is urgent to identify modifiable risk and protective factors for ASD at early age to inform prevention and intervention strategies (Zwaigenbaum et al., 2015). The pathogenesis of ASD is unknown; however, current evidence points to complex interactions between genetic and environmental factors, which represents a challenge in developing effective therapeutic approaches (Maenner, 2021, 2023). The range of genetic variants and de novo mutations associated with ASD, together with the varying number of combinations of these factors present in each individual, make it particularly challenging to target genetic factors for therapeutic approaches (Ronemus et al., 2014).

Environmental factors such as air pollution, exposure to organic toxicants, psychological stress, migration, sex, and nutritional status may have meaningful and potentially significant associations with the risk of ASD (Liu et al., 2016). Growing evidence suggests that sleep is a promising modifiable risk factor for ASD. Sleep is at the intersection of several processes consistently implicated in ASD, including neurodevelopment, immune signaling, and synaptic regulation (Karthikeyan et al., 2020). Furthermore, sleep disturbances are common in people with ASD, with up to more than half of children and adolescents reporting sleep problems (Cortese et al., 2020). A growing number of studies support a critical role for sleep in early neurodevelopmental processes. Mammals, including rodents and humans, spend significantly more time in sleep early in life, coinciding with a critical window of neurodevelopment (Alrousan et al., 2022; Wintler et al., 2020a). Sleep disruption during early life impacts a number of neurodevelopmental processes affecting sensory motor systems and social behaviors (Deliens & Peigneux, 2019; Kamara & Beauchaine, 2020). Preclinical models suggest that early life sleep disruption can cause neurobiological and behavioral alterations in adults reflective of behavioral symptoms and neurobiological factors reported in people with ASD (Mazzone et al., 2018; Wintler et al., 2020b). However, less is known about whether insufficient sleep precedes or contributes to the onset of ASD symptoms in the general pediatric population (Krakowiak et al., 2008).

Emerging research suggests that sleep disruption may be both a consequence and potential contributing factor to ASD through neurobiological mechanisms involving circadian rhythm dysregulation, melatonin pathways, and synaptic pruning abnormalities (Reynolds & Malow, 2011; Veatch et al., 2015). Furthermore, several genetic factors implicated in ASD are also involved in sleep regulation, suggesting a core genetic predisposition for sleep disturbances in people with ASD (Ji et al., 2023). While previous studies have largely focused on clinical populations with limited sample sizes (Ji et al., 2023; Kim et al., 2011), population-based studies examining age-specific sleep sufficiency in children as a potential modifiable factor associated with ASD risk remain limited.

The American Academy of Sleep Medicine (AASM) provides age-specific sleep duration recommendations to support optimal health in children (Paruthi et al., 2016b). Failure to meet these recommendations may negatively impact neurodevelopmental outcomes. Additionally, sex differences in ASD prevalence and symptom presentation have been well documented, with males approximately four times more likely to be diagnosed than females (Loomes et al., 2017; Maenner, 2021). Yet, few studies have stratified the relationship between sleep and ASD by sex and age.

Obesity is another factor associated with ASD. A greater prevalence of obesity in children with ASD has been reported (Kamal Nor et al., 2019). Sleep disturbances are often associated with obesity, including in children and adolescent populations (Cortese et al., 2020; Kamal Nor et al., 2019). Furthermore, sleep disruption is proposed to contribute to circadian rhythm, metabolic signaling, and neuroendocrine alterations that promote obesity (Kamal Nor et al., 2019). Sleep disturbances, therefore, may represent a modifiable risk factor for ASD as well as the obesity and metabolic dysfunction associated with this disorder.

Using data from a nationally representative survey, this study aims to examine the association between age-specific sufficient sleep and ASD among children aged 6–17 years in the United States. We further explore whether this relationship differs by sex, and BMI to better inform targeted sleep-based prevention strategies.

## Methods

### Study Design and Data Source

This cross-sectional study utilized data from the 2022-2023 National Survey of Children’s Health (NSCH), a U.S. Census Bureau survey sponsored by the Health Resources and Services Administration’s (HRSA) Maternal and Child Health Bureau (MCHB), according to the Census Bureau. It is the largest national and state-level survey on the health and health care needs of children ages 0-17 in the United States. The NSCH collects data via caregiver-report using web- and paper-based questionnaires and includes comprehensive information on child health, family demographics, health care access, and developmental conditions. The present analysis included children aged 6 to 17 years with complete information on ASD, sleep hours, and relevant covariates. The dataset is accessible at: https://www.census.gov/programs-surveys/nsch.html.

### Study Population

A total of 63,866 children aged 6–17 years were included after excluding individuals with missing data on ASD diagnosis, sleep sufficiency, or key covariates listed below. ASD status was determined based on the caregiver’s response to the question: *“Has a doctor or other health care provider ever told you that this child has Autism, Asperger’s disorder, pervasive developmental disorder, or ASD?”*

### Exposure Variable: Age-Specific Sufficient Sleep

The primary exposure was age-specific sleep sufficiency, defined using the American Academy of Sleep Medicine (AASM) guidelines (Paruthi et al., 2016a). Children aged 6–12 years were considered to have sufficient sleep if they slept 9–12 hours per 24-hour period, and those aged 13–18 years if they slept 8–10 hours(Paruthi et al., 2016b). Caregiver-reported usual weekday sleep duration was used to classify sleep sufficiency.

### Outcome Variable: ASD

The outcome of interest was clinical diagnosis of ASD, as reported by caregivers. This method has been validated and shown to provide reliable prevalence estimates comparable to surveillance data (Kogan et al., 2009).

### Covariates

The list of covariates included for this study were Age (continuous), Sex (male or female), Race/Ethnicity (White vs. non-White), and BMI, where BMI was classification based on age- and sex-specific percentiles: underweight (<5th), healthy weight (5th–85^th^ percentile), overweight (85th–95th), and obese (≥95th percentile), based on CDC child and teen growth charts.

### Statistical Analysis

Descriptive statistics were used to summarize population characteristics, stratified by ASD status. Continuous variables were reported as means and standard deviations (SD), and categorical variables as counts and percentages. Unadjusted and adjusted logistic regression models were used to estimate the odds ratios (ORs) and 95% confidence intervals (CIs) for the association between sleep sufficiency and ASD. Models were adjusted for age, sex, race/ethnicity, and BMI. Additionally, incidence risk ratios (IRRs), attributable incidence risk were estimated in population and risk differences (Zou, 2004). Stratified analyses by sex (male/female) were conducted to explore its potential effect on ASD modification. Interaction terms between sleep sufficiency and BMI group were tested in multivariable models. To aid interpretability and clinical translation, we also conducted machine learning prediction to quantify the potential public health impact of sufficient sleep on ASD risk profiles (Benichou, 2001). Statistical significance was set at p < 0.05. Analyses were performed using the latest version of R (version 4.5.1).

## Results

**Table 1** presents the demographics and other characteristics of the study population (N=63,866 children). Among the total sample, 4.41% (n = 2,818) were reported to have ASD, while the majority (95.59%, n = 61,048) did not. The mean age of children was 11.98 years (SD = 3.51). The sample was relatively balanced by sex, with 51.82% males and 48.18% females. The majority of participants identified as white (76.68%), while 23.32% identified as non-white.

**Table 1.** Population Characteristics of U.S. Children in the National Survey of Children’s Health (N = 63,866)

| **Characteristics** | **Levels** | **Total Subject**<br>**N = 63866 (%)** |
| --- | --- | --- |
| ASD | No | 61048 (95.59) |
|  | Yes | 2818 (4.41) |
| Age in Years | Mean (SD) | 11.98 (3.51) |
| Sex | Female | 30770 (48.18) |
|  | Male | 33096 (51.82) |
| Race/Ethnicity | non-White | 14896 (23.32) |
|  | White | 48970 (76.68) |
| BMI | Healthy weight | 39842 (62.38) |
|  | Underweight | 5449 (8.53) |
|  | Overweight | 9211 (14.42) |
|  | Obese | 9364 (14.66) |
| Age-Specific<br>Sleep Sufficiency | No | 20762 (32.51) |
|  | Yes | 43104 (67.49) |

Regarding BMI classification, most children were in the healthy weight range (62.38%), with 8.53% categorized as underweight, 14.42% overweight, and 14.66% obese. In terms of age-specific sleep sufficiency, 67.49% of children were reported to meet the recommended sleep duration guidelines based on their age, while 32.51% did not.

**Table 2** presents the prevalence of ASD across age-specific sufficient sleep and the key baseline characteristics. The mean age of children with ASD was slightly lower (11.78 years) compared to those without ASD (11.99 years). A notable sex disparity was observed: males accounted for 73.78% of ASD cases despite comprising only 51.82% of the total sample, while females represented 26.22% of ASD cases (p < 0.001), consistent with established sex-related differences in ASD prevalence. Race and ethnicity did not show a significant difference with ASD prevalence (p = 0.919), with both white and non-white children having similar proportions of ASD diagnosis. BMI classification, however, revealed significant association (p < 0.001).

**Table 2.** Prevalence of Autism Spectrum Disorder (ASD) by Baseline Characteristics Among U.S. Children Aged 6–17 Years: Estimates from the National Survey of Children’s Health (NSCH, N = 63,866).

| **Predictors** | **Total, N (%)** | **Autism Spectrum Disorder (ASD)** |  |  |
| --- | --- | --- | --- | --- |
|  |  | **Yes, N (%)** | **No, N (%)** | **p-value** |
| Total N (%): | 63866 | 2818 (4.41) | 61048 (95.59) |  |
| Age in Years: Mean (SD) | 11.98 (3.51) | 11.78 (3.48) | 11.99 (3.51) | 0.002 |
| Sex: Female | 30770 (48.18) | 739 (26.22) | 30031 (49.19) | <0.001 |
| Male | 33096 (51.82) | 2079 (73.78) | 31017 (50.81) |  |
| Race/Ethnicity: non-White | 14896 (23.32) | 660 (23.42) | 14236 (23.32) | 0.919 |
| White | 48970 (76.68) | 2158 (76.58) | 46812 (76.68) |  |
| BMI: Healthy weight | 39842 (62.38) | 1490 (52.87) | 38352 (62.82) | <0.001 |
| Underweight | 5449 (8.53) | 288 (10.22) | 5161 (8.45) |  |
| Overweight | 9211 (14.42) | 429 (15.22) | 8782 (14.39) |  |
| Obese | 9364 (14.66) | 611 (21.68) | 8753 (14.34) |  |
| Age Specified Sleep Sufficiency: No | 20762 (32.51) | 1071 (38.01) | 19691 (32.25) | <0.001 |
| Yes | 43104 (67.49) | 1747 (61.99) | 41357 (67.75) |  |

Children with ASD had a higher prevalence of obesity (21.68%) and underweight status (10.22%) compared to their peers without ASD (14.34% and 8.45%, respectively), and a lower prevalence of healthyweight (52.87% vs. 62.82%). Sleep sufficiency, defined by age-specific recommendations, was also significantly associated with ASD status (p < 0.001). A higher proportion of children with ASD had insufficient sleep (38.01%) compared to those without ASD (32.25%).

**Table 3** presents the incidence risk (IR) of ASD across age-specific sufficient sleep and demographic variables. Children with ASD were slightly younger on average than those without ASD (mean age: 11.78 vs. 11.99 years), with a modest inverse association between age and ASD risk (IR = 0.98; 95% CI: 0.91–0.99). Males had a significantly higher incidence of ASD (6.28%) compared to females (2.40%), indicating a marked sex disparity. Racial and ethnic groups showed similar ASD incidence, with no substantial difference between white (4.41%) and non-white children (4.43%). However, BMI classifications revealed higher ASD incidence among overweight (4.66%), underweight (5.29%), and obese children (6.52%) compared to those with healthy weight (3.74%). Sleep sufficiency was significantly associated with ASD risk. Children who did not meet age-specific sleep recommendations had a higher incidence of ASD (IR: 5.16, 95%CI:4.86 to 5.47) than those with sufficient sleep (IR: 4.05, 95%CI: 3.87 to 4.24) for this study population.

**Table 3:** Incidence Risk (IR) of Autism Spectrum Disorder (ASD) by Baseline Characteristics and Predictors Among Children Aged 6–17 Years: Findings from the National Survey of Children’s Health, United States (N = 63,866).

| Predictors |  | Exposure | Autism Spectrum Disorder (ASD) |  |  | Inc Risk (95% CI) |
| --- | --- | --- | --- | --- | --- | --- |
|  |  |  | ASD + | ASD - | Total |  |
| **Age in Years (Mean)** |  |  | 11.78 | 11.99 | 11.98 | 0.98 (0.91 to 0.99) |
| **Sex** | Male | Exp + | 2079 | 31017 | 33096 | 6.28 (6.02 to 6.55) |
|  | Female | Exp - | 739 | 30031 | 30770 | 2.40 (2.23 to 2.58) |
|  | Total | Total | 2818 | 61048 | 63866 | 4.41 (4.25 to 4.57) |
| **Race/ Ethnicity** | White | Exp + | 2158 | 46812 | 48970 | 4.41 (4.23 to 4.59) |
|  | non-White | Exp - | 660 | 14236 | 14896 | 4.43 (4.11 to 4.77) |
|  | Total | Total | 2818 | 61048 | 63866 | 4.41 (4.25 to 4.57) |
| **BMI** | Underweight | Exp + | 288 | 5161 | 5449 | 5.29 (4.71 to 5.91) |
|  | Healthyweight | Exp - | 1490 | 38352 | 39842 | 3.74 (3.56 to 3.93) |
|  | Total | Total | 1778 | 43513 | 45291 | 3.93 (3.75 to 4.11) |
|  | Overweight | Exp + | 429 | 8782 | 9211 | 4.66 (4.24 to 5.11) |
|  | Healthyweight | Exp - | 1490 | 38352 | 39842 | 3.74 (3.56 to 3.93) |
|  | Total | Total | 1919 | 47134 | 49053 | 3.91 (3.74 to 4.09) |
|  | Obese | Exp + | 611 | 8753 | 9364 | 6.52 (6.03 to 7.04) |
|  | Healthyweight | Exp - | 1490 | 38352 | 39842 | 3.74 (3.56 to 3.93) |
|  | Total | Total | 2101 | 47105 | 49206 | 4.27 (4.09 to 4.45) |
| **Age-Specific Sleep Sufficiency** | Yes | Exp + | 1747 | 41357 | 43104 | 4.05 (3.87 to 4.24) |
|  | No | Exp - | 1071 | 19691 | 20762 | 5.16 (4.86 to 5.47) |
|  | Total | Total | 2818 | 61048 | 63866 | 4.41 (4.25 to 4.57) |

Figure 1 illustrates the relationship between ASD and age-specific sleep sufficiency and demographic factors, using three complementary measures: Incidence Risk Ratios (IRR, Panel A), Attributable Incidence Risk (AIR, Panel B), and Attributable Incidence Risk in the Population (AIRP, Panel C). Being male gender was the strongest predictor of ASD, with an IRR of approximately 2.6, indicating that males were over 2.5 times more likely than females to be diagnosed with ASD. This sex-based difference translated to an AIR of roughly 4 additional ASD cases per 100 males and an AIRP of about 1.8 per 100 children, reflecting both high individual and population-level impacts. Children with obesity had significantly elevated ASD risk (IRR ∼1.7), corresponding to an AIR of 2.8 per 100 and an AIRP of 0.6, indicating obesity is a meaningful contributor to ASD incidence. Overweight and underweight children also had higher risks (IRRs of ∼1.25 and ∼1.4, respectively), though with smaller AIR and AIRP values. In contrast, race/ethnicity (White vs. non-White) was not significantly associated with ASD risk, showing IRR values near 1.0 and negligible AIR and AIRP estimates. Notably, age-specific sleep sufficiency emerged as a regulating behavioral factor: children with insufficient sleep had a 27% higher risk of ASD (IRR ∼1.27), an AIR of approximately 1.1 per 100, and an AIRP of 0.4.

**Figure 1:**
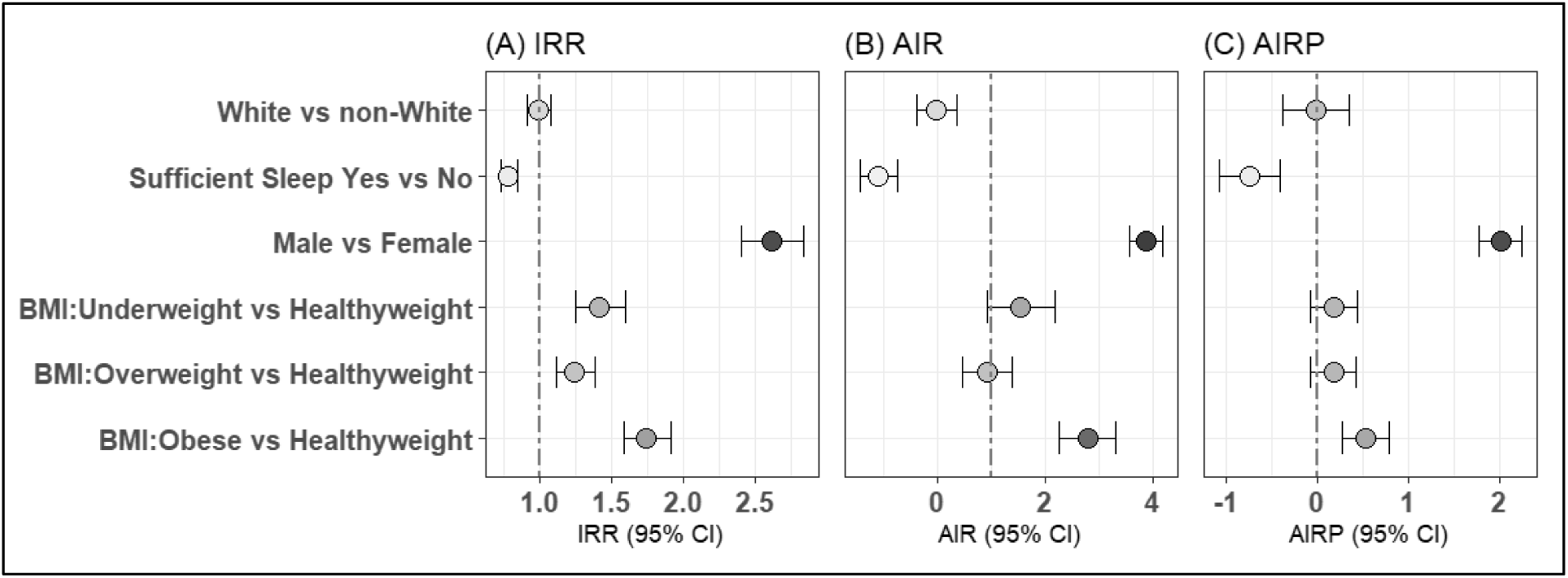
Risk of Autism Spectrum Disorder (ASD) by Baseline Characteristics and Predictors Among Children Aged 6–17 Years: Findings from the National Survey of Children’s Health, United States (N = 63,866). A) Incidence Risk Ratios, B) Attribute incidence risk, C) Attribute incidence risk to population.

Figure 2 illustrates the adjusted odds ratios (ORs) and 95% confidence intervals (CIs) for ASD based on adjusted logistic regression models. Panel A shows the overall model, while Panels B and C present sex-stratified results. In the overall analysis (Panel A), male children had significantly higher odds of ASD than females (OR = 2.66; 95% CI: 2.44–2.90; *p* < 0.001), highlighting sex as the strongest independent predictor. Children with sufficient age-specific sleep had 22% lower odds (OR = 0.78; 95% CI: 0.72–0.85; *p* < 0.001) of ASD compared to those with insufficient sleep, indicating a protective association. BMI category was also significantly associated with ASD risk: compared to children with a healthyweight, underweight (OR = 1.33; 95% CI: 1.16–1.51), overweight (OR = 1.19; 95% CI: 1.06–1.33), and obese children (OR = 1.60; 95% CI: 1.45–1.76) all had increased odds of ASD. Race/ethnicity was not significantly associated with ASD (OR = 1.03; 95% CI: 0.94–1.13; *p* = 0.48), and age was inversely associated with ASD odds (OR = 0.99; 95% CI: 0.98–1.00; *p* = 0.03).

**Figure 2.**
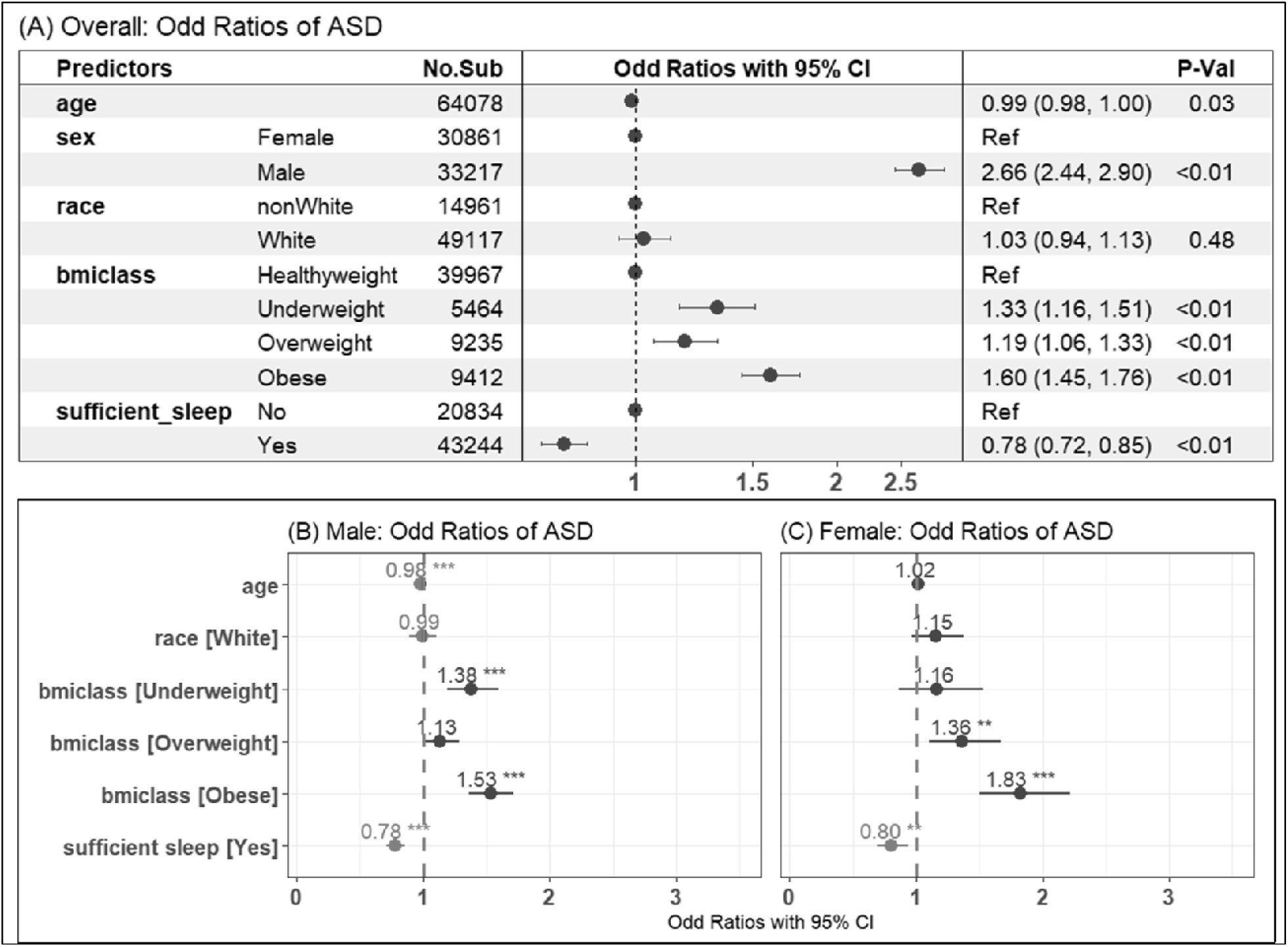
Adjusted Odds Ratios for Autism Spectrum Disorder (ASD) by Baseline Characteristics and Sleep Pattern Predictors Among Children Aged 6–17 Years: Results from the National Survey of Children’s Health, United States (N = 63,866). A) Overall logistic regression model; B) Stratified logistic regression model for males; C) Stratified logistic regression model for females.

Sex-stratified models (Panels B and C) further revealed differential effects of age-specific sleep sufficiency. Among males, obesity was associated with a 53% increase in ASD odds (OR = 1.53; 95% CI; *p* < 0.001), and both underweight and overweight statuses were also significantly associated with increased risk (ORs: 1.38 and 1.13, respectively). Sufficient sleep was protective in males (OR = 0.78; *p* < 0.001), consistent with the overall pattern. In females, the effect of obesity on ASD was even more pronounced (OR = 1.83; *p* < 0.001), and underweight and overweight statuses remained significant predictors (OR = 1.36, *p* < 0.01 for overweight). Sufficient sleep for female participants was also significantly protective (OR = 0.80; *p* < 0.01), though the magnitude of effect was slightly smaller than in males. Age showed a nonsignificant association in females, contrasting with the modest inverse association observed in males for the study population.

**Table 4** presents interaction effects of age-specific sleep sufficiency, BMI and sex in adjusted regression model with 95% CIs and p-values. A statistically significant interaction was observed between underweight and sufficient sleep (AdjOR = 0.72, 95% CI: 0.55–0.95, p = 0.018). This indicates that underweight children who received sufficient sleep had significantly lower odds of being diagnosed with ASD compared to healthyweight children who did not receive sufficient sleep. Conversely, no significant interaction effects were found (adjOR: 1.00, 95%CI: 0.79–1.25, p=0.975, for overweight, adjOR: 1.05, 95% CI: 0.86–1.28, p = 0.645, for obese) for overweight or obese children with sufficient sleep. These findings indicate that sufficient sleep did not significantly alter the odds of ASD among children that were overweight or obese. Finally, the interaction between sex and sufficient sleep was not statistically significant (AdjOR = 0.99, 95% CI: 0.83–1.18, p = 0.936), indicating that the relationship between sufficient sleep and ASD does not differ meaningfully between males and females.

**Table 4.** Interaction Effects of BMI, Sex, and Sufficient Sleep on Autism Spectrum Disorder Among U.S. Children Aged 6–17 Years: Findings from the National Survey of Children’s Health (N = 63,866).

| Predictors | ASD |  | Adj OR<br>(95% CI, P-Val) |
| --- | --- | --- | --- |
|  | No<br>N (%) | Yes<br>N (%) |  |
| **BMI:** Healthy weight | 38474 (96.3) | 1493 (3.7) | - |
| Underweight | 5176 (94.7) | 288 (5.3) | 1.66 (1.34-2.05, p<0.001) |
| Overweight | 8806 (95.4) | 429 (4.6) | 1.20 (1.00-1.43, p=0.046) |
| Obese | 8797 (93.5) | 615 (6.5) | 1.57 (1.35-1.83, p<0.001) |
| **sufficient sleep:** No | 19761 (94.8) | 1073 (5.2) | - |
| Yes | 41492 (95.9) | 1752 (4.1) | 0.81 (0.69-0.96, p=0.013) |
| **Sex:** Female | 30120 (97.6) | 741 (2.4) | - |
| Male | 31133 (93.7) | 2084 (6.3) | 2.67 (2.33-3.07, p<0.001) |
| BMI: Obese*sufficient sleep: Yes | - | - | 1.05 (0.86-1.28, p=0.645) |
| BMI: Overweight*sufficient sleep: Yes | - | - | 1.00 (0.79-1.25, p=0.975) |
| BMI: Underweight*sufficient sleep: Yes | - | - | 0.72 (0.55-0.95, p=0.018) |
| Sex: Male*sufficient sleep: Yes | - | - | 0.99 (0.83-1.18, p=0.936) |

Figure 3 presents a machine learning model used to predict the profiles of ASD risk. The most influential predictor is sex (p < 0.001), with a clear division in ASD risk between males and females. Among females, the next significant predictor is sufficient sleep (p = 0.002). For those who don’t have sufficient sleep, age is a critical factor (p < 0.001). Specifically, females aged ≤ 7 years (Node 4, *n* = 1013) have the average predicted probability of ASD close to 5%, whereas those older than 7 years (Node 5, *n* = 922) show a decreased probability, nearing 2.5%. For females who have sufficient sleep, the age threshold shifts to 14 years (p < 0.001). Females aged ≤ 14 (Node 7, *n* = 14,348) show a very low ASD risk, below 1.5%, while those older than 14 (Node 8, *n* = 6,188) show a slightly increase in risk.

**Figure 3:**
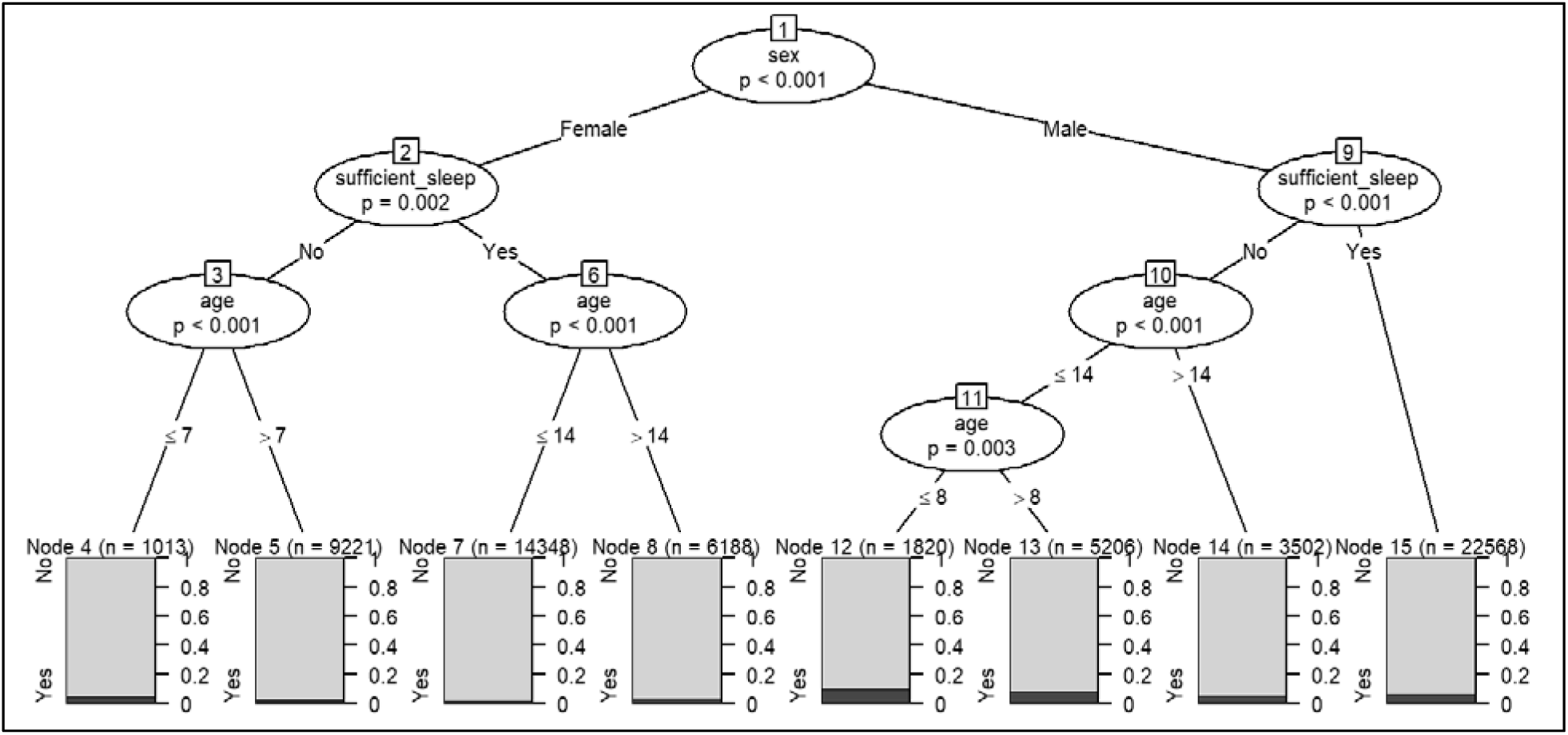
Regression Tree for predicting the probability of risk of ASD conditioning the potential predictors using baseline characteristics and age-specific sufficient sleep predictors among children aged 6–17 years, derived from the National Survey of Children’s Health (United States, N = 63,866).

In the male subgroup, sleep sufficiency again plays a crucial role (p < 0.001). Males without sufficient sleep are significantly split by age (p < 0.001). Those aged > 14 (Node 14, *n* = 3,502), display an estimated ASD probability exceeding 5%. For males aged ≤ 14, a further split occurs at age 8 years (p = 0.003). Males age 8-14 years (Node 12, *n* = 1,829) exhibit the highest ASD risk, estimated at around 12%, compared to those older than 8 but still ≤ 14 (Node 13, *n* = 5,206), whose risk increases to approximately 10%. In contrast, males with sufficient sleep (Node 15, *n* = 22,568) display lower risk among male participants, with probabilities well below 5%.

## Discussion

Our findings from a nationally representative survey data of over 63,000 U.S. children aged 6 to 17 years highlight a significant association between age-specific sufficient sleep and the risk of ASD. Children with insufficient age-specific sleep had a significantly higher risk of ASD compared to children who met the recommended sleep duration guidelines. Our findings are in line with previous studies suggesting that inadequate sleep in early childhood is linked to alterations in brain development impacting attention and social behaviors reflective of core symptoms of ASD (Reynolds & Malow, 2011; Veatch et al., 2015). Prior studies have reported that sleep problems often precede the diagnosis of ASD and may contribute to its behavioral manifestations through mechanisms such as disrupted circadian rhythms, abnormal melatonin secretion, and impaired synaptic plasticity (Cohen et al., 2014; Paruthi et al., 2016a; Veatch et al., 2015). Taken together, these findings support sleep as a modifiable risk factor for ASD.

Sufficient sleep early in life is essential for physical and mental health and brain development, and younger children require more sleep than adolescent aged children (Cohen et al., 2014; Paruthi et al., 2016b). The American Academy of Sleep Medicine (AASM) recommends 9–12 hours of sleep for children aged 6–12 years and 8–10 hours for adolescents aged 13–17 years to support their optimal health (Paruthi et al., 2016b). Our study provides further support and extends these guidelines by showing the importance of sleep in ASD risk reduction among children. This study has found that children not meeting the AASM sleep recommendations had a relative increase of more than twenty-two percent in ASD prevalence, even after adjusting for sociodemographic and health covariates.

Age was inversely associated with ASD diagnosis in our study, with younger children showing slightly higher ASD prevalence rates. This is in line with evidence for greater importance of sleep for neurodevelopment early in life, and suggests that younger children are more sensitive to the neurodevelopmental consequences due to sleep disruption (Basile et al., 2021; Cortese et al., 2020). Our findings suggest that promoting adequate sleep during childhood, particularly in early childhood, is a promising approach for reducing ASD risk.

Healthy sleeping in early life is also critical for physiological health. Decreased sleep during childhood is associated with increased risk for obesity and impaired glucose metabolism (Basile et al., 2021; Favole et al., 2023; Koren & Taveras, 2018). Short sleep duration, poor sleep quality, and sleep disruptions in children have been associated with childhood obesity (Koren & Taveras, 2018; Paavonen et al., 2009). Furthermore, insufficient sleep in children is associated with decreased insulin sensitivity and is associated with the development of metabolic disorders in both children and adults (Koren & Taveras, 2018). In our study, we found that both underweight and obese children had elevated risks of ASD compared to children in the healthy weight range. These findings are in line with previous reports for increased prevalence of obese and underweight conditions in children with ASD (Kamal Nor et al., 2019, 2019; Koren & Taveras, 2018). Furthermore, this is in agreement with growing evidence suggesting that nutritional status and metabolic dysregulation may contribute to neurodevelopmental disorders through inflammatory, endocrine, and gut-brain axis pathways (Dash et al., 2022; He et al., 2025).

Sex-stratified models revealed that the association between insufficient sleep and ASD was more pronounced in males, who were already more likely to be diagnosed with ASD. This is consistent with the longstanding epidemiologic trend of higher ASD prevalence in boys (male-to-female ratio ∼4:1), potentially due to a combination of biological susceptibility, diagnostic bias, and protective factors in females such as hormonal and genetic mechanisms (Lai et al., 2015; Loomes et al., 2017). Importantly, while females had a lower overall ASD risk, insufficient sleep still emerged as a notable risk factor, highlighting the relevance of adequate sleep for both sexes. No significant differences in ASD prevalence were observed between White and non-White children after adjusting for other variables. While disparities in diagnosis rates across racial and ethnic groups have been documented in prior research, often driven by access to care and cultural perceptions, recent national efforts may be reducing this gap in diagnosis rates (Pham & Charles, 2023; B. Zhang et al., 2024).

Current evidence suggests that decreased sleep during childhood may arise from both genetic and environmental factors. For example, several genetic factors implicated in ASD, such as Fmr1, Nlgn3, Shank3, Cacna1c, and Mef2c are also involved in sleep regulation (Doldur-Balli et al., 2022; Y. Zhang et al., 2024). Thus, the presence of one or more of these genetic factors may contribute to sleep disturbances early in life and, in turn, to neurodevelopmental and metabolic dysfunction. Maternal immune activation and two-hit immune activation models, commonly used to model neurodevelopmental dysfunction associated with ASD, result in male specific sleep and circadian rhythm alterations in offspring (Hall et al., 2023; Kang et al., 2024). Future studies examining the presence of sleep disturbances together with genetic and behavior factors may serve as a promising approach for identifying at risk children and allow for age-specific sleep based therapeutic and preventative strategies to alleviate the severity of ASD symptoms early in the disease process (Alhowikan et al., 2019; Kang et al., 2024).

## Public Health Implications

Our results have critical implications for early intervention and prevention. While genetic predisposition plays a substantial role in ASD, modifiable lifestyle factors such as adequate sleep and healthy weight maintenance can offer practical, non-invasive avenues for risk reduction.

Integrating sleep health education into pediatric care and school-based programs may help identify at-risk children and support healthy developmental trajectories. Furthermore, promoting public awareness of the risks of insufficient sleep for childhood health and guidelines for sleep hygiene habits for children, such as proper environmental temperature, noise exposure, and ambient light, regular sleep routines and sleep-wake schedules, and proper timing of exercise, meals, and screen time may assist in improving sleep duration and quality. A growing number of studies indicate that early school start times are associated with sleep problems in children, and that later start times are associated with decreased sleepiness along with improved developmental outcomes and mood. This study contributes to the growing public health literature on ASD and insufficient sleep among children, underscoring the urgent need for continued research and the development of effective intervention strategies to address this critical issue.

## Technical Limitations

Key strengths of this study include its large, nationally representative sample, the use of standardized and age-specific sleep criteria, and stratified analyses that reveal sex-specific patterns. However, several limitations must be acknowledged. First, ASD diagnosis and sleep duration were caregiver-reported, introducing the potential for recall and reporting bias. Second, the cross-sectional design limits causal inference. Future longitudinal studies are needed to clarify temporal relationships between sleep insufficiency and ASD onset. Third, residual confounding from unmeasured variables (e.g., family history, screen time, comorbidities) may remain. Lastly, our dataset did not include actigraphy or EEG measures to allow for analysis of sleep architecture and circadian rhythms. Future studies incorporating these measures in large cohorts would be needed to identify which aspects of sleep are associated with ASD risk, such as REM sleep, NREM sleep, circadian period length and amplitude.

## Conclusion

This study provides robust population-level evidence that insufficient age-specific sleep is significantly associated with elevated ASD risk, with variation across sex, age, and BMI status. In addition, females under the age of 14 with sufficient sleep exhibited the lowest likelihood of ASD, while males aged 8 to 14 years with insufficient sleep showed the highest risk of ASD. These findings underscore the importance of exploring age-specific sleep sufficiency strategies early in life as a potential strategy to reduce neurodevelopmental vulnerability, particularly among high-risk groups. Longitudinal and interventional research is warranted to further evaluate sleep as a target for early ASD risk mitigation.

## Statements and Declarations Supplementary Information

None

## Data Availability

The data from the study are available on National Survey of Children’s Health (NSCH) website (https://www.census.gov/programs-surveys/nsch.html)

## Declarations

### Ethical Approval

This study did not involve any procedures requiring ethical approval.

### Informed Consent

Not applicable, as the study utilized publicly available, de-identified data.

## Acknowledgments

We extend our deepest appreciation to NSCH for giving us access to the dataset.

